# Variable-Selection ANOVA Simultaneous Component Analysis (VASCA)

**DOI:** 10.1101/2022.06.13.22276334

**Authors:** José Camacho, Raffaele Vitale, David Morales-Jimenez, Carolina Gómez-Llorente

## Abstract

**Motivation:** ANOVA Simultaneous Component Analysis (ASCA) is a popular method for the analysis of multivariate data yielded by designed experiments. Meaningful associations between factors/interactions of the experimental design and measured variables in the data set are typically identified via significance testing, with permutation tests being the standard go-to choice. However, in settings with large numbers of variables, the “holistic” testing approach of ASCA (all variables considered) often overlooks statistically significant effects encoded by only a few variables.

**Results:** We propose Variable-selection ASCA (VASCA), a method that generalizes ASCA through variable selection, augmenting its statistical power without inflating the Type-I error risk. The method is evaluated with simulations and with a real data set from a multi-omic clinical experiment. We show that VASCA is more powerful than both ASCA and the widely-adopted False Discovery Rate (FDR) controlling procedure; the latter is used as a benchmark for variable selection based on multiple significance testing. We further illustrate the usefulness of VASCA for exploratory data analysis in comparison to the popular Partial Least Squares Discriminant Analysis (PLS-DA) method and its sparse counterpart (sPLS-DA).

**Availability:** The code for VASCA is available in the MEDA Toolbox at https://github.com/josecamachop/MEDA-Toolbox

**Supplementary information:** Supplementary data are available at *Bioinformatics* online.

## 1 Introduction

Designed experiments control the variation of one or more *factors* to assess the effect of such variation on one or more specific responses (variables) of interest. As an example, imagine that blood samples are extracted from two cohorts of cancer patients treated with two distinct drugs and characterized through chromatographic measures; one may then want to understand if and how the chromatographic profile of these blood specimens changes when altering the therapeutic strategy (*i*.*e*., the type of drug). If a single response is considered, the classical approach to identify those factors (e.g., drug type) significantly effecting the observed variation is Fisher’s analysis of variance (ANOVA) (Fisher, 1918). To assess the significance of such effect, a given test statistic (e.g., the F ratio) is used to compute Fisher’s *p*-value, which is then compared with certain threshold for statistical significance. For multivariate responses (current state of the art in, *e*.*g*., omics sciences like genomics, transcriptomics, proteomics or metabolomics), a widely adopted approach is to conduct multiple (univariate) tests over the individual variables, in an attempt to identify and select the specific responses significantly affected by a given experimental factor. A univariate test statistic (as in ANOVA) and corresponding *p*-value is therefore obtained for each variable. Statistical significance thresholds in such multiple testing are typically adjusted (corrected) in order to identify as many significant variables as possible, while keeping the number of false positives under control according to some criterion; well-known examples are the Bonferroni correction to control the family-wise error rate (FWER), and the Benjamini-Hochberg (BH) and Benjamini-Yekutieli (BY) procedures (Benjamini and Hochberg, 1995; Benjamini and Yekutieli, 2001) to control the False Discovery Rate (FDR). FDR-controlling procedures (or simply, *FDR methods*) are particularly appealing in multivariate settings with large numbers of variables due to their increased statistical power—they allow to identify more significant associations due to the less conservative *p*-value correction compared to e.g., the Bonferroni corrrection. Thus, FDR methods—and particularly the BH procedure—have been widely used for multiple significance testing in omics data. However, as they are based on univariate tests, FDR methods do not take into account the possibly complex multivariate structure underlying the data. Indeed, the effect of an experimental factor may be reflected as certain inter-dependencies or combinations of several measured variables (e.g., a linear combination of responses), rather than simply as variation within individual variables. This limits the detection power of the FDR approach.

An alternative strategy to treat multiple responses is to apply multivariate testing procedures, such as Multivariate ANOVA (MANOVA) (Warne, 2014). These methods replace multiple univariate tests by a single multivariate one to assess the statistical significance of the ANOVA model, including all the variables in the data set. This avoids the need for multiple-testing corrections, but suffers from the effects of statistical noise, particularly when the number of variables is large, leading to a remarkable lack of discriminatory power. The multivariate test statistic often contains noisy contributions from a large number of (insignificant) variables, and meaningful associations of factors affecting only a few variables can potentially be buried within the overall statistical noise.

ANOVA Simultaneous Component Analysis (ASCA Smilde *et al*. (2005); Jansen *et al*. (2005)) is one popular multivariate extension of ANOVA, widely employed in, e.g., chemistry, biology and biomedicine (Smilde *et al*., 2005; Nueda *et al*., 2007; Bevilacqua *et al*., 2013; De Luca *et al*., 2016; Du *et al*., 2017; Firmani *et al*., 2020). It combines the variance factorization and inference capabilities of ANOVA with the exploratory power of Principal Component Analysis (PCA). In ASCA, the statistical significance of factors’ and interactions’ effects is typically estimated by permutation testing (Anderson and Braak, 2003; Vis *et al*., 2007): basically, the data variation induced by such effects is contrasted against an empirical null-distribution obtained by resampling. Albeit this strategy shows notable advantages compared to other testing approaches (Vis *et al*., 2007), it inherits the limitations of multivariate testing. Indeed, the “holistic” testing approach of ASCA (all variables considered) often fails to find any statistical significance for factors associated with only a reduced sub-set of variables.

In this article, we propose a generalization of ASCA by introducing a new method for variable selection, termed Variable-selection ASCA (VASCA). The main idea is to incorporate variable selection in the multivariate permutation testing procedure of ASCA to robustly assess the statistical significance of the experimental model. The proposed testing procedure attains improved detection power without compromising the Type-I error risk, while still being able to fully capture the inherent multivariate nature of the data. The enhanced statistical power brought by variable selection leads to improved ASCA modelling; for a given effect (factor/interaction), VASCA identifies significant associations with a selected sub-set of variables, filtering out those not accounting for the said effect, and narrowing down the subsequent ASCA analysis to a reduced amount of meaningful responses.

VASCA is here assessed with simulations and with a real data set from a multi-omic clinical experiment, and compared to ASCA and the BH (FDR) method in terms of statistical power, and to Partial Least Squares Discriminant Analysis (PLS-DA) and its sparse counterpart (sPLS-DA) in terms of exploratory power. We also include a comparison with an early approach for variable selection in ASCA, the ASCA-genes method by Nueda *et al*. (2007).

## 2 ANOVA Simultaneous Component Analysis (ASCA)

Following the idea of ANOVA, ASCA is based on several steps: (1) factorization of the data according to the factors/interactions of the experimental design; (2) significance testing (based on permutation tests) for factors/interactions; (3) visualization of significant factors/interactions using Principal Component Analysis (PCA) to understand separability among levels; and, optionally, (4) post-hoc testing of levels using confidence intervals.

### 2.1 Factorization of the data

Let **X** be the *N* × *M* data matrix collected in a designed experiment. For simplicity and without loss of generality, we will take the example of a design with two fixed factors. The data in **X** can be decomposed as

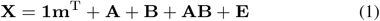

where **1** is a vector of ones of suitable length, **m** represents the overall mean (i.e., a vector with the means of all the measured variables), **A** and **B** represent the factor matrices, **AB** the interaction matrix and **E** the residual matrix. The idea behind this decomposition is to partition the variance observed in the data set **X**, according to the different factors/interactions of the experiment. In this paper, we use the technique referred to as ASCA+ (Thiel *et al*., 2017) to account for mild unbalancedness in the data. Basically, the decomposition is derived as the least squares solution of a regression problem, where **X** is regressed onto a coding scheme **C** built from the experimental design:

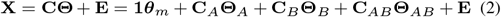

where **C** = [**1, C**_*A*_, **C**_*B*_, **C**_*AB*_] is defined following Thiel *et al*. (2017) and **Θ** = [***θ***_*m*_, **Θ**_*A*_, **Θ**_*B*_, **Θ**_*AB*_] and **E** are obtained as

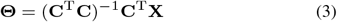

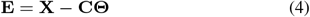

Thus, the factorized data associated with a given factor/interaction *Z* is computed as **Z** = **C**_*Z*_ **Θ**_*Z*_. This solution minimizes the variance in the residual matrix **E**.

### 2.2 Statistical significance testing

Permutation testing in the context of ASCA can be performed by randomly shuffling the rows of **X** in Equation (3), yielding a new set of regression coefficients:

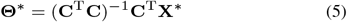

where superscript ∗ stands for permuted. Then, the permuted factorized data for any factor/interaction *Z* is re-computed as 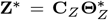 and the error as **E**^∗^ = **X**^∗^ − **CΘ**^∗^. Equivalently, one can permute the rows or values in **C** instead of those in **X** (Camacho *et al*., 2022).

Permutation tests are carried out by comparing a given statistic, computed from the ASCA-factorized data set, with the corresponding statistic computed from hundreds or more permutations. The *p*-value is obtained as^1^

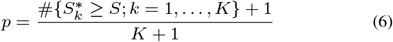

where *S* refers to the statistic computed from the factorized data matrix, 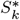 is the statistic corresponding to the *k*-th random permutation, and *K* is the total number of permutations. Thus, the *p*-value yields an empirical estimate of the probability of obtaining a result as (or more) extreme as the observed one when the null hypothesis holds.

There are several choices for the ASCA test statistic—see Camacho *et al*. (2022) for a recent review on the permutation approach and the relevance of the chosen statistic. The (Type-I) sum-of-squares of the factor 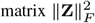 was proposed as the original one (Vis *et al*., 2007). Motivated by the data visualization aspect of ASCA, which typically uses the first 2 Principal Components (PCs), Zwanenburg *et al*. (2011) proposed testing the sum-of-squares of the first 2 PCs of the factor matrix. This is also used by Thiel *et al*. (2017). More recent variants of ASCA (Marini *et al*., 2015; Martin and Govaerts, 2020) employ the F-ratio, computed as the ratio of the mean sum-of-squares of the factor/interaction and the suitable next order factor/interaction (often the residuals). Finally, in ASCA extensions for unbalanced data (Thiel *et al*., 2017; Martin and Govaerts, 2020), the Type-III sum-of-squares is proposed; this is computed from the difference between the residuals in the reduced and full models (see references above for more detail).

### 2.3 Visualization and Post-hoc tests

Several visualization methods have been proposed that combine the ANOVA-like decomposition in Equation (2) with subspace visualization, in particular with PCA. Among these, ASCA and ANOVA-PCA (APCA) are closely related (Zwanenburg *et al*., 2011; Thiel *et al*., 2017) and both share the same approach in the factorization and inference (significance testing) steps, prior to visualization. For visualization and post-hoc testing, APCA performs PCA on each significant factor/interaction matrix plus the unexplained variance: **Z** + **E**. ASCA, however, carries out PCA on **Z**, and then visualizes the projection of **Z** + **E** in the corresponding score plot. In both approaches, the score-plot is used for the visualization and possible significance testing (Liland *et al*., 2018) of the differences among levels. In Figure 1, a simulated example of visualization with confidence intervals is presented.

**Fig. 1:**
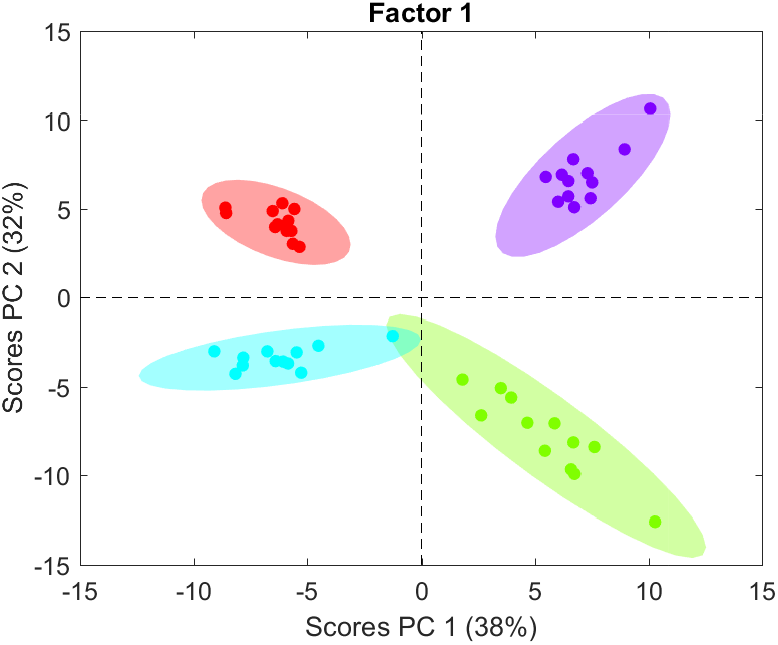
Example of ASCA visualization with confidence intervals in artificial data.

Since this paper is focused on the statistical significance testing aspect of ASCA (permutation testing), the proposed solution can be applied to both ASCA(+) and APCA(+), but we will focus only on the former.

## 3 VASCA

VASCA is based on breaking down the single test statistic computed for significance testing in ASCA (all variables considered), i.e., *S* in eq. (6), into variable-wise statistics *S*^*v*^ for *v* ∈ {1, ‥, *M*}. This derivation is straightforward for any choice of the statistic (see previous discussion in Section 2.2) based on the sum-of-squares, since this can be computed for each variable independently, and it holds that *S* = ∑_*v*_ *S*^*v*^. Since the F ratio is also based on (mean) sum-of-squares, the computation of variable-wise F ratios is also direct, but the previous equality does not necessarily hold. For the rest of the paper we will refer to the standard test statistic of ASCA, i.e., the Frobenius norm (sum-of-squares) of the factorized data 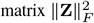.

VASCA starts by sorting out the variables in decreasing order of *S*^*v*^. Then, it iteratively assesses statistical significance of the data matrix composed of the first *m* variables in this ordering, with *m* ranging from 1 to the total number of variables *M*. The null-hypothesis is rejected for the largest significant data matrix, and the subset of selected variables therein are called significant. In this regard, VASCA is at the same time i) a multivariate version of the step-up BH (FDR-controlling) procedure (Benjamini and Hochberg, 1995) and ii) a generalization of ASCA, since VASCA includes the assessment of significance for the entire data set with *M* variables. Therefore, VASCA is at least as powerful as ASCA.

Let us define:

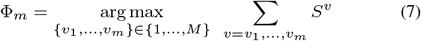

as the sub-set of *m* variables Φ_*m*_ = {*v*_1_, …, *v*_*m*_} that maximizes the sum statistic 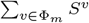. We refer to 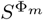 as the corresponding multivariate statistic for this sub-set, i.e., that computed from the whole data (sub)matrix rather than from individual variables. For *S* defined as the Frobenius norm (sum-of-squares) of a factorized data matrix, it holds:

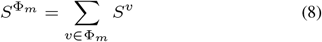

Testing for significance in VASCA requires careful consideration of the permutations in the selected variables. This is key to provide a meaningful null-distribution and to guarantee the statistical power of the approach. Rather than using multiple-testing corrections like in the BH procedure, we embed the variable-selection mechanism within the permutation testing. Within each permutation *k* we reorder the variables in decreasing order of the permuted, variable-wise statistic 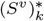. For a given number of variables *m*, the sub-set of variables in the permutation is recomputed as 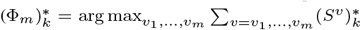, so that selected variables can (and will most likely) be different to those in Φ_*m*_. Then, we recompute the corresponding statistic 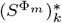. The set of statistics 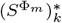 for different permutations *k* (and sub-sets of *m* variables) characterizes the null-distribution and is employed to compute the *p*-value for statistical significance:

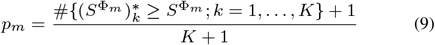

This means that in any given sub-set of *m* variables, significance is assessed by contrasting the true 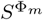 with the permuted 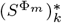, where the specific *m* variables may not be the same. For some statistics, this may require some form of normalization. In this paper we always use 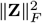 as the test statistic, and we auto-scale (normalize to 0 mean and unit variance) the data so that further normalization is not necessary.

Given that we select the maximum *m* for which significance is found, i.e., max(*m*) with *p*_*m*_ below the significance threshold, and due to the multivariate nature of VASCA, (sub)matrices called significant may indeed include some non-significant variables. This is due to the filtering nature of multivariate models and may be seen as a disadvantage over univariate tests such as BH (FDR). However, from the set of *m* selected variables, a subsequent analysis based on PCA can help distinguishing variables that are truly relevant from those that are not. To this end, we propose a bootstrapping procedure to identify PCA loadings (variables) with statistically significant values—i.e., significantly different to 0. Thus, all variables corresponding to non-significant loadings are discarded.

### 3.1 Connection with the ASCA genes method

The closest approach to VASCA is the ASCA-genes method by Nueda *et al*. (2007). ASCA-genes identifies relevant variables with permutation testing, but with two main differences in comparison to VASCA. First, the main statistic under analysis in ASCA-genes is the PCA model leverage; the residuals, measured by the squared prediction error (SPE), are also used to differentiate among types of variables. Most relevant variables are expected to present a high leverage and low SPE. Variables with high SPE and leverage are considered as poorly modelled but still potentially interesting. High SPE variables with low leverage are considered odd. Variables with low leverage and low SPE are considered not relevant. Rather than splitting the variance of the model into leverage and SPE, VASCA makes use of the complete sum-of-squares (or a similar statistic) in the factorized matrices, which simplifies interpretation, is more flexible (e.g., can be applied to models with 1 component) and is a more established method for inference in ASCA. It is also more consistent with the methodology of ANOVA: if the factor/interaction is not significant, the comparison between levels (in this case with PCA), which often implies multiple comparisons, is not performed. Second, and most importantly, ASCA-genes is based on a regular permutation approach, while VASCA is grounded on the reordering of variables within each permutation. This extra precaution is key to avoid false positives.

## 4 Evaluation in simulation examples

We devised four simulated experiments in order to compare VASCA with ASCA and the BH procedure to control the FDR, simply referred to as FDR from now on. For simplicity and for the sake of clarity, in the first three examples we simulate a single factor with two levels, where each level includes 40 subjects (thus a total of 80) for which 400 variables are collected. This choice is motivated by the nature of typical omics experiments (Tenorio-Jiménez *et al*., 2019). In the last example, we consider a two factor/multi-level problem.

To simulate the background in the multivariate data **X**, we used the SimuleMV tool (Camacho, 2017). SimuleMV allows to simulate a data set with a certain level of correlation. The inputs to SimuleMV are the size of the data matrix (number of rows and columns) and a level of correlation between 0 (absence) and 10 (maximum correlation). In our simulations, we chose level 7. Each simulated experiment is repeated 100 times and average and standard deviation results are presented.

For comparison purposes, FDR was implemented from the (empirical) probability distribution obtained through permutation testing, and corrected *p*-values (through the BH procedure) were computed. In all cases, the permutations used the same seed for the random generation engine in the three methods, ASCA, FDR and VASCA. P-values are computed with 1000 permutations.

### 4.1 Example 1: Non-significant relationship

The first example illustrates the case where the data matrix **X** and the class coding **C** for the factor are unrelated. We generate **X** with the SimuleMV tool. **C** is obtained in two steps: first, we draw 80 observations *l*_*i*_ from a normal distribution with zero mean and standard deviation one; second, we assign each of the 80 rows of **X** to one of the classes depending on the sign of the corresponding observation *l*_*i*_, and we construct **C** following that assignment. We repeat this data generation procedure 100 times. Given the independent generation of **X** and **C**, we expect no statistical significance to be found in the analysis by any of the methods.

Results are shown in Figure S1, which depicts the ordered *p*-values obtained for FDR and VASCA, and the single *p*-value (for the complete matrix **X**) for ASCA. Average results are shown with the corresponding lines and the shadowed areas represent standard deviations. Typically considered thresholds for statistical significance at 0.05 and 0.01 are also included as control limits. Note that vertical axes are in logarithmic scale.

We can see that all methods yield *p*-values well above the control limits, illustrating their robustness against Type-I errors. We can also see that VASCA generally matches ASCA in this example. Note the exact match at 400 variables, as expected. Similar results are obtained for **X** drawn at random from a multinormal distribution.

### 4.2 Example 2: Significant one-to-one relationships

We start by generating **C** and **X** as in the previous example. Then, we modify three variables in **X** so that a significant bias is induced between observations from the two classes.The simulation is designed to generate a one-to-one relationship between each of the three variables and the simulated factor in **C**. We expect both the FDR method and VASCA to identify these relationships, but because they are only present in 3 out of the 400 variables, we expect ASCA to overlook it.

Results are presented in Figure 2. As expected, FDR and VASCA systematically identify the one-to-one relationships (*p*-value < 0.001 for the first variables), while ASCA identifies the factor as non-significant, well above the control limits. Again, we can see that VASCA exactly matches ASCA for the complete set of 400 variables, since the former generalizes the latter. This example illustrates the variable selection capability of VASCA as a clear advantage over ASCA.

**Fig. 2:**
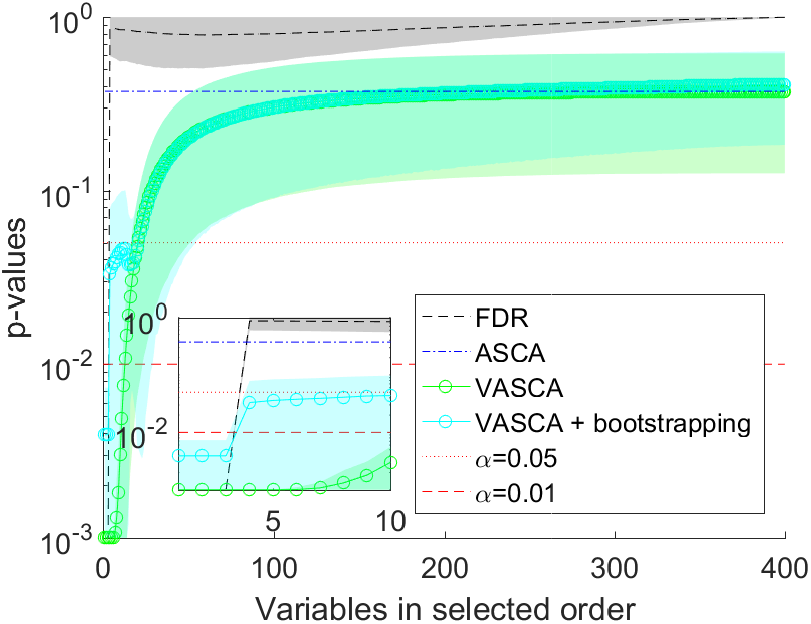
One-to-one relationships between 3 variables in **X** and **C**. Comparison of *p*-values computed with FDR, ASCA and VASCA (with bootstrapping).

The zoomed image in Figure 2 reflects that VASCA identifies models up to 6-10 variables as significant (*p*-value < 0.001 for 6 variables, *p*-value < 0.01 for 10). Contrarily, FDR accurately identifies only three variables as truly significant. The outcome of VASCA is a consequence of the filtering nature of multivariate models, i.e., models with 6/10 variables are called significant, even when only 3 of these variables are truly associated with the factor. Significance results are however refined in the subsequent analysis step of VASCA. To see this, let us proceed with the workflow of VASCA, by visualizing the factorized data with PCA in Figure S2. The scores, shown in panel A, clearly distinguish the two classes, confirming the significance of the model^2^. The loadings in panel B show that 3 variables are by far the most relevant of the 6 under study (shown by the magnitude of the loadings). The bootstrapping intervals indicate that only those three variables are statistically significant (loadings significantly different from 0). We applied the same bootstrap approach to the 100 repetitions in the simulation, to depict a curve of significance for VASCA + bootstrapping in Figure 2, showing the same accuracy as the FDR method at a *p*-value threshold of 0.01.

Finally, we wanted to check what would happen if we repeat the same simulation scheme of the example, but with a much smaller bias in the three variables, so that the variance that reflects the connection between them and the factor is 10 times smaller than in the previous case. The result is shown in Figure S3 We can see that the FDR does not detect a significant relationship on average anymore. VASCA, however, shows a higher statistical power and determines on average a p-value < 0.05 for the most significant variable. This higher power is the result of VASCA taking advantage of the correlation between the three variables associated to the factor.

### 4.3 Example 3: Multivariate relationship

In this example, we generate **X** with the SimuleMV tool except for three variables that are drawn from a normal distribution with mean zero and standard deviation one. We thus make these three variables independent from the rest and no spurious correlation is created between **C** and **X**. Again, **C** is obtained in two steps: we first obtain 80 values *l*_*i*_ by summing the three normal variables in **X**; then, we assign each of the 80 rows of **X** to one of the design levels depending on the sign of the corresponding sum, and we build **C** following that assignment. With this approach, we have created an additive multivariate relationship between the three variables in **X** and **C**. Therefore, unlike the previous example, we need to consider the combination of the three variables to properly differentiate the aforementioned levels. Additive multivariate relationships are consistent with the interpretation of biomarkers in networks of pathways, where correlations are identified as paths that jointly contribute to a response/reaction.

Results are presented in Figure 3. Overall, the FDR method does not detect significance in this situation, which was expected due to its univariate nature. VASCA is generally more powerful, increasing the chances to detect significance, which on average is only found at *p*-value < 0.05 and for two variables (see the zoomed detail in the figure). This illustrates the complexity behind capturing multiple additive (possibly cancelling) effects in high-dimensional data. The ulterior bootstrapping in VASCA does not reduce the statistical power. ASCA once again overlooks the relationship. This example illustrates the benefits brought by the multivariate nature of VASCA which, as opposed to the FDR method, is able to identify multivariate additive relationships among variables.

**Fig. 3:**
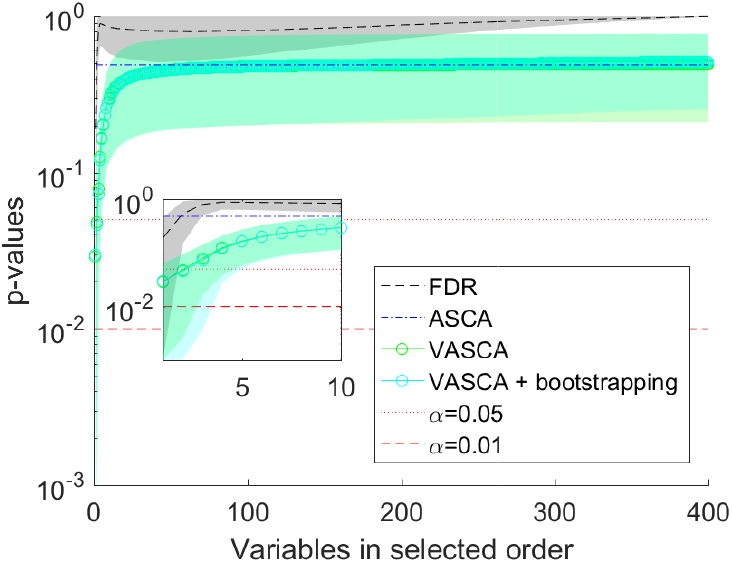
Multivariate relationship between 3 variables in **X** and **C**. Comparison of *p*-values computed with FDR, ASCA and VASCA.

### 4.4 Example 4: Multivariate relationship in two factors and interaction with several levels

A major strength of ANOVA, also inherited by ASCA, VASCA or FDR, is the analysis of complex experimental designs with several factors and interactions. In this last experiment we simulate data sets with two significant factors, of four and three levels, respectively. Each factor is simulated as in the previous example, so that a significant multivariate relationship exists between the factor and three variables.

Results are presented in Figure S4. To some extent, they resemble those in the previous example, with an increased power of VASCA in comparison to FDR. However, in this case significance was not systematically found: we can see that average results of the 100 simulation repetitions are above the 0.05 significance level. This example illustrates the cumbersome issues that may arise when trying to recognise particular effects influencing multivariate data coming from complex experimental designs. The methods did nonetheless find significance in a sub-set of the 100 repetitions, as shown in Table 1, where VASCA outperforms ASCA and FDR in the number of positive identifications. We also see that the use of bootstrapping can reduce the inferential power of the approach— meaning that the bootstrapping test is more conservative than VASCA’s permutation testing. This should be taken into account when permutation testing and bootstrap yield conflicting results, e.g., for datasets with only 1 significant variable found by VASCA and then deemed as non-significant by bootstrap. For instance, take the example of factor 1. Out of the 100 simulations, VASCA finds 37 with at least 1 significant variable and 20 with at least 2. Therefore, it finds exactly 17 (which comes from subtracting 37 minus 20) simulations with only 1 significant variable. When using bootstrap, this number is reduced to only 9 cases (3 subtracted by 12).

**Table 1.**
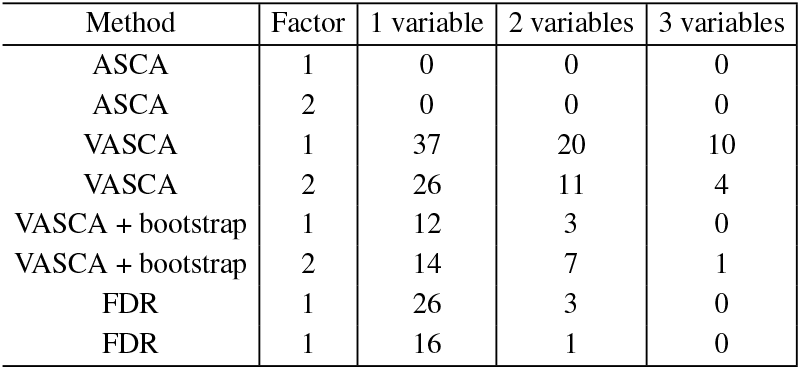
Number of simulations (out of the total of 100) where at least 1, 2 and 3 variables were found statistically significant in the example with the multivariate relationship between 3 variables in **X** and **C** (2 factors of 4 and 3 levels, respectively, are taken into account here). Comparison of FDR, ASCA and VASCA (with and without bootstrapping).

| Method | Factor | 1 variable | 2 variables | 3 variables |
| --- | --- | --- | --- | --- |
| ASCA | 1 | 0 | 0 | 0 |
| ASCA | 2 | 0 | 0 | 0 |
| VASCA | 1 | 37 | 20 | 10 |
| VASCA | 2 | 26 | 11 | 4 |
| VASCA + bootstrap | 1 | 12 | 3 | 0 |
| VASCA + bootstrap | 2 | 14 | 7 | 1 |
| FDR | 1 | 26 | 3 | 0 |
| FDR | 1 | 16 | 1 | 0 |

For further comparison, we illustrate the result obtained by the ASCA-genes method by Nueda *et al*. (2007) in one randomly selected simulation. To compute both leverage and residuals, the PCA model corresponding to a factor in ASCA needs at least 2 PCs, and thus the factor needs to include at least 3 levels. ASCA-genes is thus not applicable to the examples described in sections 4.1 to 4.3. Results obtained with ASCA-genes in the 2-factor (4 and 3 levels) example of the present section can be found in Figure S5. Two of the significant variables (marked in dark color) for Factor 1 show high leverage. The other significant variable is found in the SPE chart. For factor 2, however, there are several non-significant variables that exceed the leverage control limit, which should be regarded as false positives. In this specific example, VASCA detected two significant variables for Factor 1 (the third presented a *p*-value close to 0.05) and none for Factor 2, FDR could only detect one significant variable for Factor 1, and VASCA + bootstrap none. We can see that VASCA and ASCA-genes yield similar results, with VASCA being more general (i.e., it can be applied to rank-one factor/interaction matrices) and less prone to false positives.

## 5 Results on real data

The BIOASMA data set (Gomez-Llorente *et al*., 2020) comprises clinical, biochemical, anthropometrical parameters, inflammatory biomarkers, metagenomic and metabolomic data for 46 children (12 girls and 34 boys, aged 4-13 years) with an allergic asthma diagnosed based on the Spanish Guidelines for Asthma Management (GEMA criteria 4.4)— (Moral *et al*., 2016). The children were also classified into normal-weight (n=13), overweight (n=8) and obese (n=25) according to the age and sex-specific thresholds proposed by Cole *et al*. (2000). Biochemical data were obtained by routine methods. Inflammatory biomarkers were determined by ELISA and by XMAP Luminex technology. Metabolomic data were obtained by one-dimensional proton nuclear magnetic resonance (1D ^1^H-NMR) spectra of blood plasma samples. Short chain fatty acids were determined by Gas Chromatography-Mass Spectrometry. Metagenomic data were obtained by 16sRNA barcoding sequencing and the Amplicon sequence variants (ASVs) were normalized by the rarefaction method. Deriving potential biomarkers from this data set represents a real challenge (Gomez-Llorente *et al*., 2020), given the low sample size and the complexity of the experimental design: two potential conflicting factors (asthma severity and weight classification/status) with three levels each are taken into account and the individuals distribution is significantly unbalanced.

### 5.1 Factor-wise models

The statistical results obtained from the original analysis of the data set (Gomez-Llorente *et al*., 2020) are based on Partial Least Squares Discriminant Analysis (PLS-DA) (Barker and Rayens, 2003) and its sparse variant sPLS-DA (Lê Cao *et al*., 2008). sPLS-DA considers variable selection during model calibration with the idea of discarding non-informative variables. Neither PLS-DA nor sPLS-DA models were statistically significant to distinguish the three classes of weight status or the three classes of asthma severity. However, for individual sPLS-DA models for both factors it was possible to find statistically significant differences between one of the classes vs the rest. In particular, a sPLS-DA model with 12 variables was found statistically significant to distinguish the persistent asthma class from the rest (occasional and frequent asthma) with an Area Under the Receiver Operating Characteristics curve (AUROC) of 0.66 ±0.08 (p-value < 0.05) in double cross-validation (Szymańska *et al*., 2012), and a sPLS-DA model with three variables was found statistically significant to distinguish the normo-weight class from the rest (overweight and obese) with an AUROC of 0.75 ± 0.09 (p-value < 0.05). The PLS-DA version of the first model (asthma severity) with only selected variables is presented in Figure S6, where the scores show a clear separation between persistent asthma and the rest. The model for weight status is discussed in the Supplementary Material.

We analyze the asthma severity in Figure 4 following the same approach as in the simulated data, i.e., we compare the ordered *p*-values obtained for FDR and VASCA, and the single p-value for ASCA. Control limits highlighting significance for a *p*-value < 0.05 and a *p*-value < 0.01 are also shown, and the vertical axes are in logarithmic scale. Figure S7 illustrates the results when we consider the three classes (occasional, frequent and persistent asthma), and Figure 4 when we consider persistent asthma vs the rest. In both situations, ASCA is in agreement with PLS-DA showing no statistical significance. VASCA is in agreement with sPLS-DA and significance is only found for a sub-set of variables when two classes (persistent asthma vs the rest) are considered. The FDR method fails to find statistical significance, arguably as a consequence of not directly accounting for the multivariate nature of the investigated data.

**Fig. 4:**
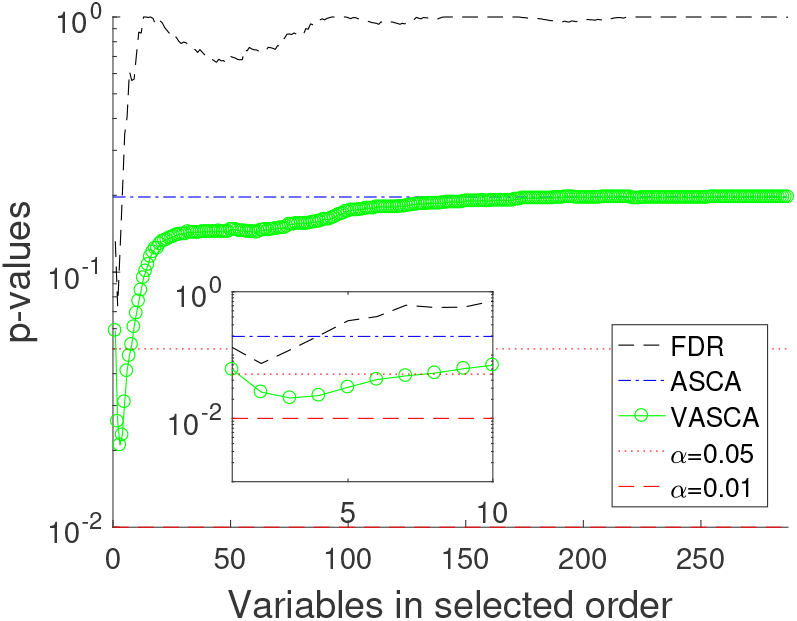
Comparison of *p*-values computed with FDR, ASCA and VASCA for the BIOASMA data set (persistent asthma vs the rest).

Figure 5 shows the scores and loadings for the single component in the VASCA model, which should be compared to the corresponding sPLS-DA biplots in Figure S6. In the VASCA model, all variables present loadings which are significantly different to 0 (p-value < 0.05). Looking at the scores, both sPLS-DA and VASCA show similar separation ability, but VASCA selects only half of the analyzed variables, those in the left part of Figure S6.

**Fig. 5:**
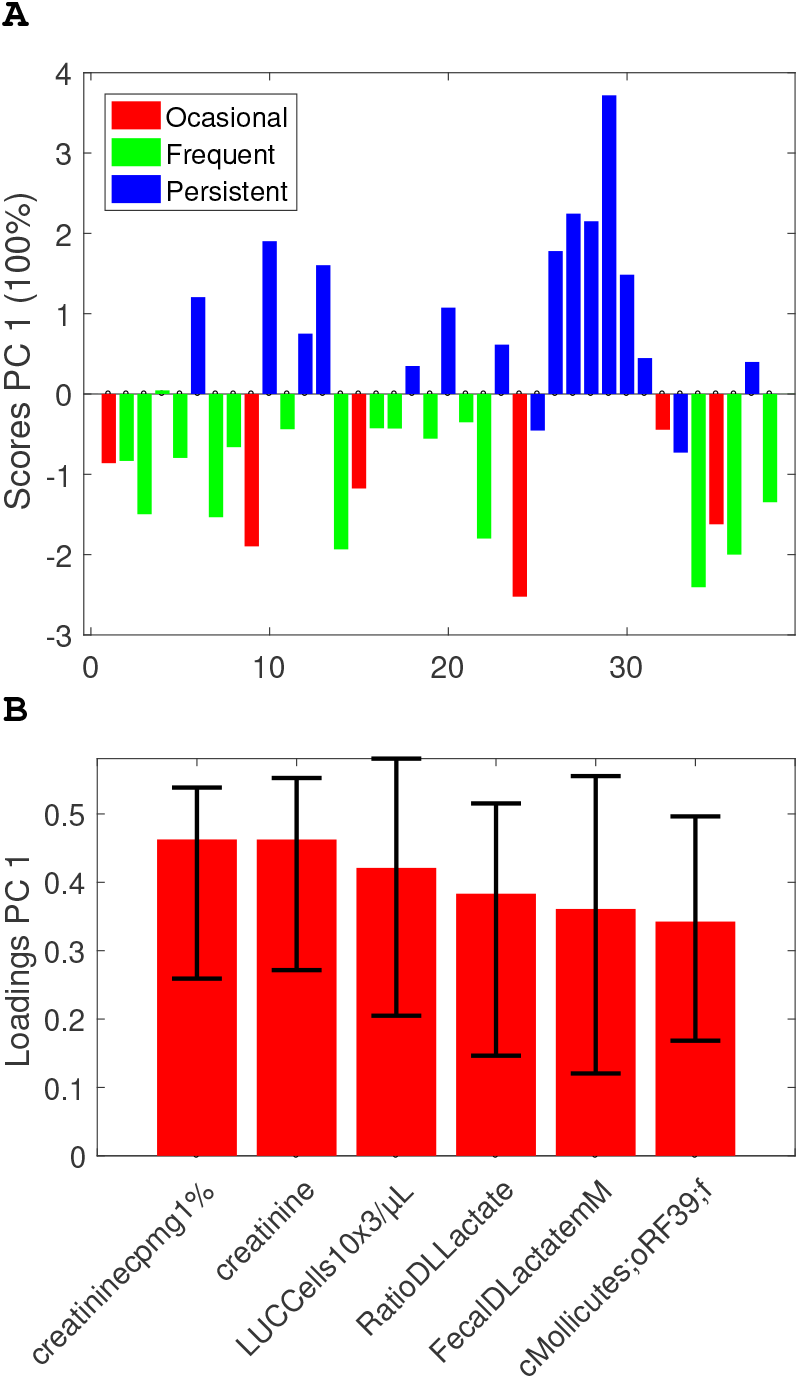
VASCA (6 variables) scores (a) and loadings (b) plots for the BIOASMA data set (persistent asthma vs the rest).

To provide a numerical assessment of the variable selection by sPLS-DA and VASCA, we compute the AUROC of the reduced PLS-DA models constructed on the variables selected by each method. Note that these AUROC values are expected to be overoptimistic, since they are computed from the same data used for variable selection. Yet, they are useful to compare the ability of both methods (sPLS-DA and VASCA) to find potentially interesting biomarkers. We obtain 0.99 ± 0.01 for sPLS-DA with 12 selected variables and 0.93±0.02 for VASCA with 6 selected variables.

The same analysis with the weight factor (normo-weight, overweight and obese) can be found in the Supplementary Material (see Figures S8 and S9). In this case, ASCA does not find any significant model (just like PLS-DA) while both FDR and VASCA find a single significant variable. We obtain 0.73±0.02 for PLS-DA with the 3 variables selected by sPLS-DA (Gomez-Llorente *et al*., 2020) and 0.74 ± 0.01 for PLS-DA with the individual variable selected by VASCA/FDR. This shows that, beyond its inference and variable selection capabilities, VASCA can be a competitive exploratory tool for, e.g., biomarker identification.

### 5.2 Multi-factor models

In this section we consider simultaneously the two factors of the BIOASMA data set in a single analysis. Figure S10 shows the comparison of *p*-values for ASCA, FDR and VASCA for two levels in each factor: persistent asthma vs the rest (occasional and frequent asthma) and normo-weight vs the rest (overweight and obese). No statistically significant results were obtained for the three levels in any of the factors. For two factors and two levels (one class vs the rest), only VASCA found statistically significant results. Interestingly, in the VASCA model the second factor (the weight classification) is not significant anymore and only the first factor is. This comes as a result of the ANOVA factorization of both factors together, and shows that separated PLS-DA models can lead to the double counting of variance that can hamper interpretation of the results.

## 6 Conclusion

In this paper, we presented VASCA, an extension of ANOVA Simultaneous Component Analysis (ASCA) that improves the statistical inference of multivariate models through variable selection. VASCA is inspired by the popular Bejamini-Hochberg (BH) step-up procedure to control the False Discovery Rate (FDR). Its benefits are two-fold: first, by taking on the idea of variable selection from FDR-controlling procedures, it attains substantially improved discrimination (detection) power over conventional ASCA; and second, based on multivariate inference (similar to ASCA), it is able to model/capture and visualize inherent multivariate relationships within the experimental data. Our results showed that VASCA can outperform both the BH (FDR-controlling) procedure and ASCA in terms of statistical power, and that it represents a competitive exploratory approach in comparison to widely used techniques such as Partial Least Squares Discriminant Analysis and its sparse counterpart.

## Data Availability

No real data is explicitly generated for this paper. The code for the simulation study is available upon reasonable request to the authors. The code for VASCA is available in the MEDA Toolbox at
https://github.com/josecamachop/MEDA-Toolbox

## Footnotes

1 Please, note in the equation we assume that the higher the statistic the more significant. We will maintain this assumption during the rest of the paper.

2 Unlike Figure 1 that shows a score plot with two PCs, the present example encompasses a factor of two levels which makes the factorized matrix in ASCA of rank one, and only one PC can be extracted.

