## Supplementary Materials for "Variable-Selection ANOVA Simultaneous Component Analysis (VASCA)"

### 1. Evaluation in simulation examples

#### *1.1. Example 1: Non-significant relationship*

The results of this experiment are shown in Figure S1. The figure presents the ordered $p$-values obtained by FDR and VASCA, and the single $p$-value (for the complete matrix $\mathbf{X}$) returned by ASCA. Average results are shown with the corresponding lines and the shadowed areas represent standard deviations. Control limits highlighting significance for a $p$-value $< 0.05$ and a $p$-value $< 0.01$ are also displayed. Note the vertical axes are in logarithmic scale. The figure shows that all methods yield $p$-values well above the control limits, illustrating their robustness against Type-I errors.

#### *1.2. Example 2: Significant one-to-one relationships*

Scores and loadings of the VASCA model are shown in Figure S2. The scores (left) clearly distinguish the two design levels confirming the significance of the model. The loadings (right) show that 3 variables are by far the most relevant of the 6 under study. If we derive bootstrapping intervals, we can see that only the loadings for those three variables are significantly different from 0.

The result of the One-to-one relationships between 3 variables in $\mathbf{X}$ and $\mathbf{C}$ created with a small bias is shown in Figure S3. We can see that the FDR does not detect a significant relationship on average anymore. VASCA, however, shows a higher statistical power and determines on average a p-value $< 0.05$ for the most significant variable.

[1]Signal Theory, Networking and Communications Department, University of Granada, C/ Periodista Daniel Saucedo Aranda s/n 18071, Granada, Spain

[2]Univ. Lille, CNRS, LASIRE (UMR 8516), Laboratoire Avancé de Spectroscopie pour les Interactions, la Réactivité et l'Environnement, F-59000, Lille, France

[3]Department of Biochemistry and Molecular Biolgoy II, School of Pharmacy, Institute of Nutrition and Food Technology "José Mataix", Biomedical Research Center, University of Granada 18160, Granada, Spain. ibs.GRANADA, Instituto de Investigación Sanitaria, 18012, Granada, Spain. CIBEROBN (Physiopathology of Obesity and Nutrition CB12/03/30038), Instituto de Salud Carlos III, 28029, Madrid, Spain

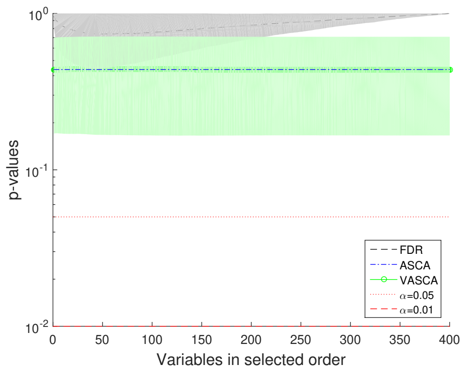


Figure S1: No relationship between **X** and **C**. Comparison of $p$-values computed with FDR, ASCA and VASCA.

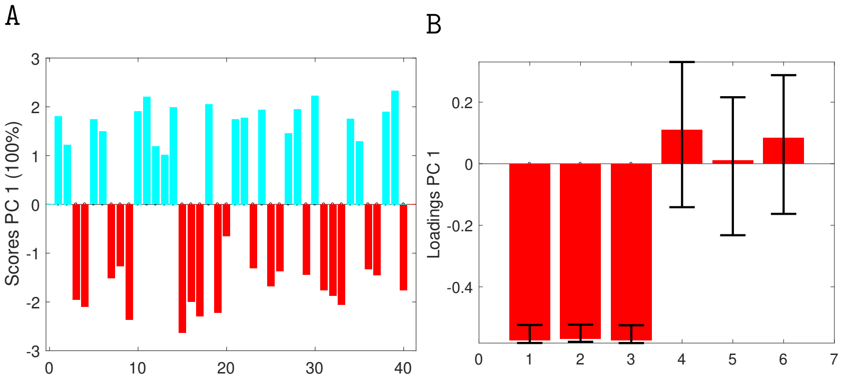


Figure S2: VASCA (6 variables) scores (a) and loadings (b) plots. Simulation with one-to-one relationships between 3 variables in **X** and **C**.

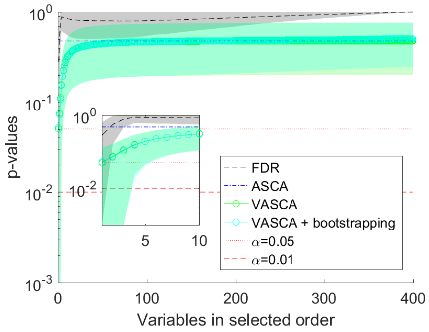


Figure S3: One-to-one relationships between 3 variables in **X** and **C** created with a smaller bias. Comparison of $p$-values computed with FDR, ASCA and VASCA (with bootstrapping).

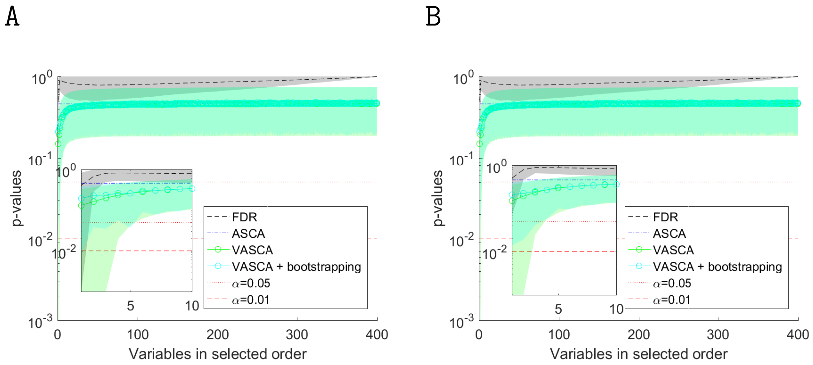


Figure S4: Multivariate relationship between 3 variables in **X** and **C** (2 factors of 4 and 3 levels, respectively, are taken into account here). Comparison of $p$-values computed with FDR, ASCA and VASCA (with bootstrapping). Factor 1 (a) and 2 (b)

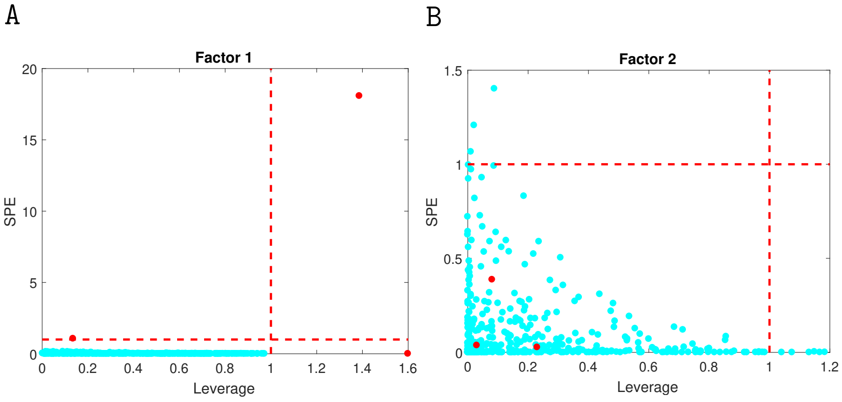


Figure S5: Variable selection by the ASCA-genes method. Factor 1 (a) and 2 (b). Ground-truth significant variables are represented in dark (red) color.

#### *1.3. Example 4: Multivariate relationship in two factors and interaction with several levels*

The comparison results are presented in Figure S4. To some extent, the outcomes resemble those of Example 3 in the main manuscript, with an increased power of VASCA in comparison to FDR.

We illustrate the results yielded by the ASCA-genes method [2] in Figure S5. Two of the relevant variables (marked in dark red color) varying with Factor 1 show high leverage. The other significant variable is found in the SPE chart. For factor 2, however, there are several non-significant variables that exceed the leverage control limit, which should be regarded as false positives. In this specific example, VASCA detected two significant variables for Factor 1 (the third presented a $p$-value close to 0.05) and none for Factor 2, FDR could only detect one significant variable for Factor 1, and VASCA + bootstrap none.

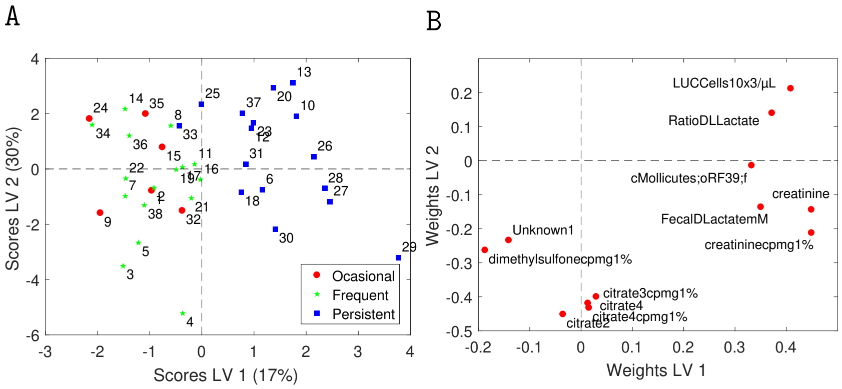


Figure S6: PLS-DA model for class "persistent asthma" vs the rest (occasional and frequent asthma) in the BIOASMA data set: (a) scores plot and (b) loadings plot.

### 2. Results on real data

#### 2.1. *Model for Asthma severity*

A sPLS-DA model with 12 variables was found statistically significant [1] to distinguish the persistent asthma class from the rest (occasional and frequent asthma) with an Area Under the Receiver Operating Characteristics curve (AUROC) of $0.66 \pm 0.08$ (p-value $< 0.05$) in double cross-validation [3], and a sPLS-DA model with three variables was found statistically significant to distinguish the normo-weight class from the rest (overweight and obese) with an AUROC of $0.75 \pm 0.09$ (p-value $< 0.05$). The PLS-DA model for asthma severity with only selected variables is presented in Figure S6, where the scores show the clear separation between persistent asthma and the rest.

We analyze the asthma severity in Figure S7 following the same approach as for the simulated data, comparing the ordered $p$-values resulting from FDR and VASCA, and the single $p$-value returned by ASCA. Control limits highlighting significance for a $p$-value $< 0.05$ and a $p$-value $< 0.01$ are also displayed, and the vertical axes are in logarithmic scale. Figure S7 illustrates the results when we consider the three classes (occasional, frequent and persistent asthma).

#### 2.2. *Model for Weight classification*

The PLS-DA model for weight classification with only selected variables is presented in Figure S8, where the scores show the separation between normo-weight and the rest.

We analyze the weight classification in Figure S9 following the same approach as before. Figure S9(a) illustrates the results when we consider the three classes (normo-weight, overweight and obese), and Figure S9(b) when we consider normo-weight vs the rest. In both situations, ASCA is in agreement with PLS-DA showing no statistical significance. VASCA and FDR are in agreement with sPLS-DA and significance is only found for a sub-set of variables when two classes (normo-weight class vs the rest) are considered.

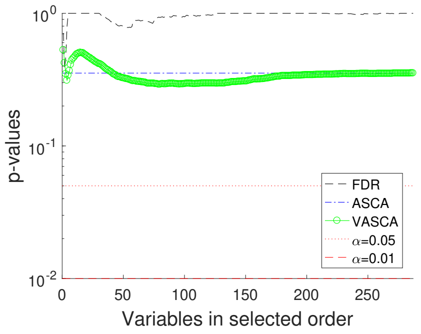


Figure S7: Comparison of $p$-values computed with FDR, ASCA and VASCA for the BIOASMA data set (occasional, frequent and persistent asthma.

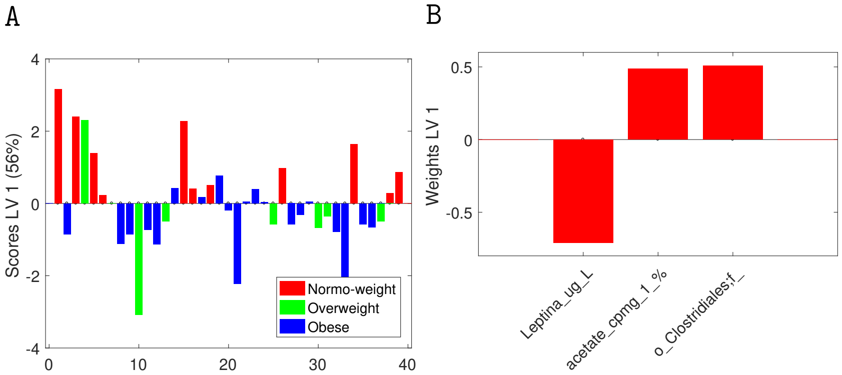


Figure S8: PLS-DA model for class "persistent asthma" vs the rest (occasional and frequent asthma) in the BIOASMA data set: (a) scores plot and (b) loadings plot.

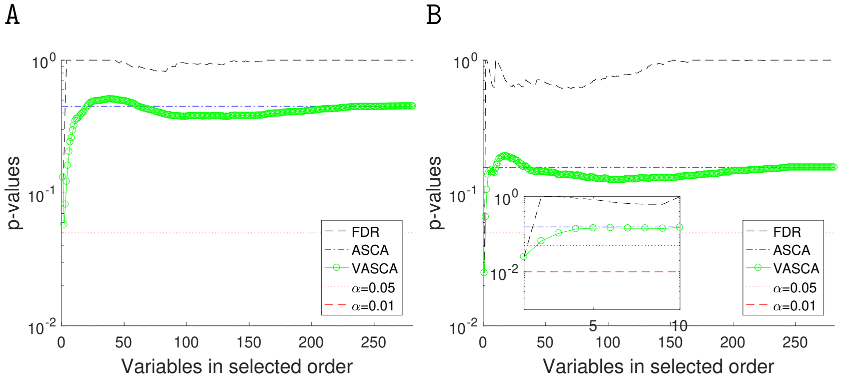


Figure S9: Comparison of $p$-values computed with FDR, ASCA and VASCA for the BIOASMA data set – (a) normo-weight vs overweight vs obese and (b) normo-weight vs the rest.

*2.3. Multi-factor model*

In this section we consider simultaneously the two factors of the BIOASMA data set in a single analysis. Figure S10 shows the comparison of $p$-values for ASCA, FDR and VASCA taking into account two levels within each factor: persistent asthma vs the rest (occasional and frequent asthma) and normo-weight vs the rest (overweight and obese). No significant results were obtained when all three levels are considered within the two factors.

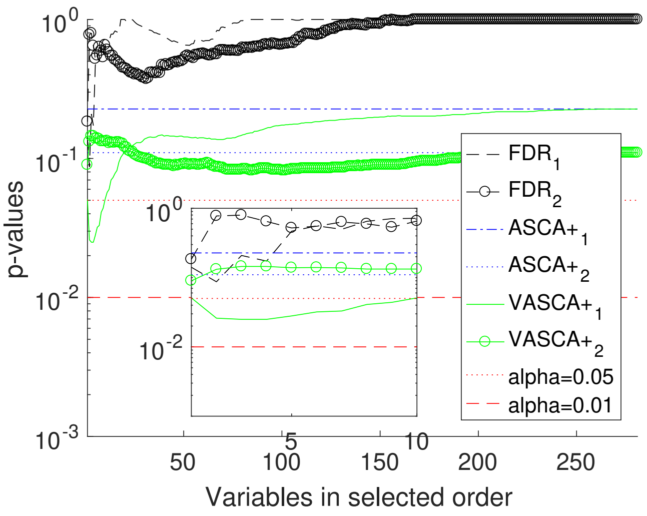


Figure S10: Comparison of $p$-values computed with FDR, ASCA and VASCA.
